# Compassion training influences state but not trait level heart-rate variability within severe depression

**DOI:** 10.1101/2023.01.10.23284408

**Authors:** Jeffrey J. Kim, Chase Sherwell, James N. Kirby

## Abstract

Heart-rate variability (HRV) is a marker of parasympathetic nervous system activity, and is a robust predicter of improved mental and physical health. Current psychotherapeutic interventions are effective at reducing self-report depressive symptoms, but few have improved HRV within a sample of severe depressive symptoms. This study explores the impact of a brief Compassion Focused Therapy exercise (CFT) on HRV. Results suggested that a brief CFT exercise can successfully target depressive physiology, at two distinct timepoints, pre- and post-a two-week self-directed training period. Specifically, we first show that CFT can significantly improve HRV at the state level, but not at the trait level after a two-week intervention. Second, CFT can increase a subset of participants’ HRV above a clinical cut-off of low resting HRV. Third, the frequency of practice (i.e., listening to the audio recording) during the self-directed training component was very low, with 50% not accessing the practice. Finally, during the CFT exercise at post-two-week training HRV decreased across time, indicating participants had a greater engagement in the ‘threat’ component of the CFT exercise – a feature to be more fully examined in prospective studies. This study suggests the value of future research with larger-scale randomized control trials, to further explore the modulation of parasympathetic physiology using compassion practices.

## Introduction

Individuals at risk for mental and physical health concerns have lower heart-rate variability (HRV)^1–4^, a marker of parasympathetic nervous system activity^5,6,7^. One specific population that have been found to exhibit lower HRV are those with depression^8^. Depression is one of the most common mental health disorders worldwide^9^, and meta-analyses have demonstrated those with major depression have lower HRV compared to healthy populations^10,11^. Although a range of therapy programs (e.g., cognitive behavior therapy^12^, acceptance and commitment therapy^13^, and mindfulness based therapies^14^) have shown efficacy in reducing self-report depressive symptoms, little work has examined training programs that can increase HRV^5,6^. Importantly, while cognitive behaviour therapy and mindfulness based cognitive therapy have successfully reduced self-reported depressive symptoms, these training modalities have not been able to shift HRV^15–17^.

In contrast Compassion Focused Therapy (CFT) seeks to actively increase HRV^18–21^. Based on CFT’s theoretical model of social mentality theory, individuals at risk of mental health problems in times of threat are often reliant on competitive social mentalities, which are linked to a physiological pattern textured by dominant sympathetic and low parasympathetic system regulation^19^. In turn, this reliance on competitive motives increases susceptibility to self-criticism, shame, and depressive symptomatology^20^, which is associated with lower HRV^21^. A core aim of CFT is to strengthen the individual’s internal physiological regulation systems, such that there is an improved parasympathetic response, indexed by improved HRV. There is strong meta-analytic evidence that greater levels of compassion are associated with increased HRV (Hedges *g* = .54)^22^, and that actively generating compassion toward others can also increase HRV^21^. Moreover, researchers have found that brief CFT interventions can increase HRV, but these studies are typically conducted with university student populations and not with those with severely depressed symptoms^5^. One successful approach that has been applied to those with depression is using HRV biofeedback, which a meta-analysis found a medium effect size (*g* = .38) at improving HRV, which had a positive impact on reducing depressive symptomatology^23^. The aim of this study was to determine whether a brief self-directed CFT training could improve HRV in those with severe depressive symptomatology.

The CFT exercise we used was a key technique in the therapy called ‘The Compassionate-Self’. In CFT, this technique is central to the therapeutic process, as the compassionate-self is actively cultivated so that the individual can then work with their self-criticism, shame or other emotional difficulties with this compassionate orientation^5-6^. The compassionate-self is typically completed as an imagery-based guided exercise and includes three core elements: 1) grounding and body posture, 2) soothing-rhythm breathing, and 3) cultivating the compassionate-self (specifically cultivating wisdom, strength and commitment to be compassionate). Once, the person has then embodied this compassionate-self orientation, we then apply the compassionate-self to work with the individual’s self-criticism. At this stage in the exercise the participants are instructed to imagine something they are self-critical about, and then asked as their compassionate-self imagine what they could say, what the feel towards, and would like to do for this part of themselves experiencing the self-criticism. In this way, the exercise has an imagery behavioural rehearsal component, where the individual imagines what they could do to compassionately help the part of themselves that feels criticised. Previous research has identified that this compassionate-self exercise central to CFT can increase HRV within a healthy population^24–26^, and reduce symptoms of depression, anxiety, and stress, and fears of self-compassion^27^. Importantly, no work has yet to demonstrate these effects within a sample of elevated depression symptomatology

Although research has positioned CFT as an effective treatment option for depression^19,26,28–30^, no work has yet to provide compelling evidence that CFT exercises can improve HRV within depression^31,32^. In the present paper, therefore, we offer an investigation of a core CFT exercise, ‘the compassionate-self’ within a sample elevated in depression. Specifically, we replicated a design used with healthy controls that examined HRV response during CFT pre and post a two-week self-directed training period^5,32^. Based on CFT’s theoretical model and research published previously with healthy controls, we first expected that CFT exercises will significantly increase HRV compared to a resting state^5,23,32–34^. Second, we expected that how often participants engaged in the compassionate-self practice across the two-week period will be a significant factor in differential HRV response^32,35^. Third, we expected to shift a subset of participants above a clinical cut-off of low-resting HRV^3,32^. Fourth, we will assess whether we could alter trait-level HRV, by assessing the baseline HRV after a two-week training period, given previous work has suggested an increase in baseline HRV within a control sample^5^. In doing so, we will examine how HRV may be modulated by CFT within a sample of elevated depression symptomatology, and provide proof-of-concept evidence to support CFT’s recommendation to target depression via modulation of parasympathetic physiology.

## Methods

### Participants

41 participants (32 Female, 9 Male; Mean age = 22.51, SD = 3.26) with elevated depression symptomatology (Mean Depression = 21.24, SD = 10.36; Mean Anxiety = 25.00, SD = 9.14; Mean Stress = 16.28, SD = 8.33) took part in the present study. Based on the Depression Anxiety and Stress Scales – Short Form (DASS-21) clinical cut-offs^36,^ the scores on the Depressive subscale placed our population in the severely depressed range and the severely anxious range for the anxious subscale. The first author’s University Human Research Ethics Committee approved the experimental protocol, and all participants provided written and/or digital informed, voluntary consent, and received remuneration at the value of $40 Australian dollars for participation.

### Measures

This study used the Short Form of the Depression Anxiety Stress Scale (DASS-21^36^) to derive a marker of elevated depressive symptomatology. Items are scored on a scale from 0 (Did not apply to me at all) to 3 (Applied to me very much, or most of the time) across the three subscales: depression, anxiety, and stress. Here, higher scores indicate greater levels of symptomatology. An example item from the measure includes “I found it difficult to relax”. The DASS has been found to have excellent internal consistency for the combined subscales at α□=□0.88^37^. Whilst it is an option to score all three subscales separately or as a total score, we used the Depression subscale, which in our sample was shown to have an excellent internal consistency of α□= 0.89.

### HRV Design

Participants listened to a 15-minute recording of the CFT exercise “Cultivating the Compassionate Self”, with HRV analysed during a baseline recording and during the CFT meditation exercise. This was the same exercise recorded pre- and post-a two-week self-directed engagement period (Figure 1). As mentioned, the “*Cultivating the Compassionate Self*” exercise has three core elements; 1) grounding and body posture, 2) soothing-rhythm breathing, and 3) cultivating the compassionate self. Importantly, during the third core element, ‘cultivating the compassionate self’, individuals are asked to bring to mind a current mistake, setback or failure in their life, which they are self-critical of, and to orient their compassion-self toward this. Details on this meditation have been reported previously and validated within a healthy control sample^32^, and materials and instructions for accessing the website can be found on the Open Science Framework (OSF) (10.17605/OSF.IO/PWZGJ). The guided exercise was recorded in English by a Clinical Psychologist and expert in CFT. A website to facilitate this project during the two-week engagement period between Time 1 and Time 2 was created to house the audio recording and various instructions/helpful tip sheets for participants (https://exp.psy.uq.edu.au/self/index.html).

**Figure 1.**
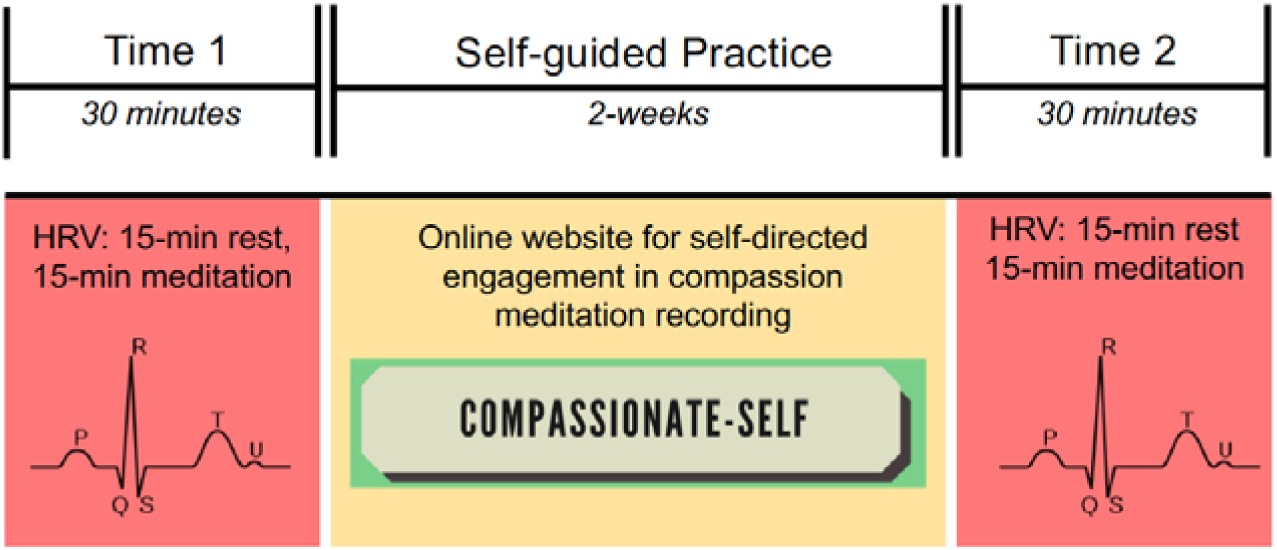
Task diagram for current experiment. A 15-minute Rest and 15-minute Compassion Meditation HRV recording was collected at each timepoint. Separating each timepoint was a 2-week self-guided practice, with an online website for self-directed engagement in the same audio-recording participants listened to at each timepoint.

### HRV data acquisition, pre-processing

We collected our heart-rate variability (HRV) data with a portable Schiller Systems Medilog electrocardiogram (ECG) device. These recorders are the “AR12plus” type and use a 3-lead bipolar ECG amplifier. We positioned each lead on the chest, to maximize ECG amplitude and minimize muscle artifacts. After data collection, data were digitized, pre-processed, and extracted using Schiller’s proprietary Medilog Darwin software package, which processes the ECG biosignal to comply with accepted guidelines in the field for measurement and analysis of HRV data. Via visual inspection of the ECG biosignal, and after application of an algorithm which automatically filters for normal and non-normal beats, we excluded signal artifacts and irregular (non-normal) beats not removed from the initial application of the pre-processing algorithm. Therefore, only the normal beats (via R-wave peak detection) were extracted for the HRV analysis. The sampling period was 1000 Hz at a time resolution of 250 μs (microseconds), and we utilized linear detrending.

After initial pre-processing, high-frequency (HF) HRV components (i.e., 0.15-0.40 Hz) of each participants’ ECG trace were extracted upon application of the discrete Fourier transform (DFT) of the beat-to-beat interval times-series. We also extracted raw RMSSD HRV components, namely the Root Mean Square of Successive Differences of the HRV beat-to-beat timeseries. Data were coded as being from either Time 1 or Time 2, and at either Rest or Intervention. Means for these periods were created from the bins downloaded from Medilog Darwin (i.e., 4*15-minute, 12*5-minute, and 60*1-minute bins, across Rest and Meditation, at both Time 1 and Time 2). It should be noted that we did not measure or control for respiration during the ECG recording, yet we did attempt to control for posture, exercise, alcohol, caffeine intake, and time of day effects. HF HRV data were transformed with the natural logarithm in order to better approximate normality^38,39^, whereas RMSSD HRV data remained in the ‘raw’ scale to examine elevated log ratios (below). Across the two weeks, participants engaged with the meditation on average 3.5 times (*SD* = 3.45, Range = 0-12 listens).

## Results

### High-Frequency (HF) HRV Response

We conducted ANOVAs on our 15-minute binned HRV data, to examine overall group differences in means. First, we conducted an ANOVA with the normalised HF data. Here, a two-way ANOVA revealed a significant effect of session (i.e., rest vs intervention), whereby HRV was higher when engaged in the compassion intervention than at rest (*F*(1,38) = 15.04, *p* <.001). We also observed a non-significant effect of time (i.e., pre and post two-week training) (*F*(1,38) = 1.30, *p* = .26), whereby rest versus intervention did not differ between timepoints, as well as a non-significant interaction term between these two factors (*F*(1,38) = .03, *p* = .88). Follow-up t-tests revealed significant increases in HRV from T1 rest to T1 intervention (t(38) = 3.34, *p* <.002), as well as from T2 rest to T2 intervention (t(40) = 2.88, *p* <.006). An overall t-test from T1 rest to T2 intervention was non-significant (t(39) = 1.17, *p* = .251). HF-HRV response is shown in Figure 2.

**Figure 2.**
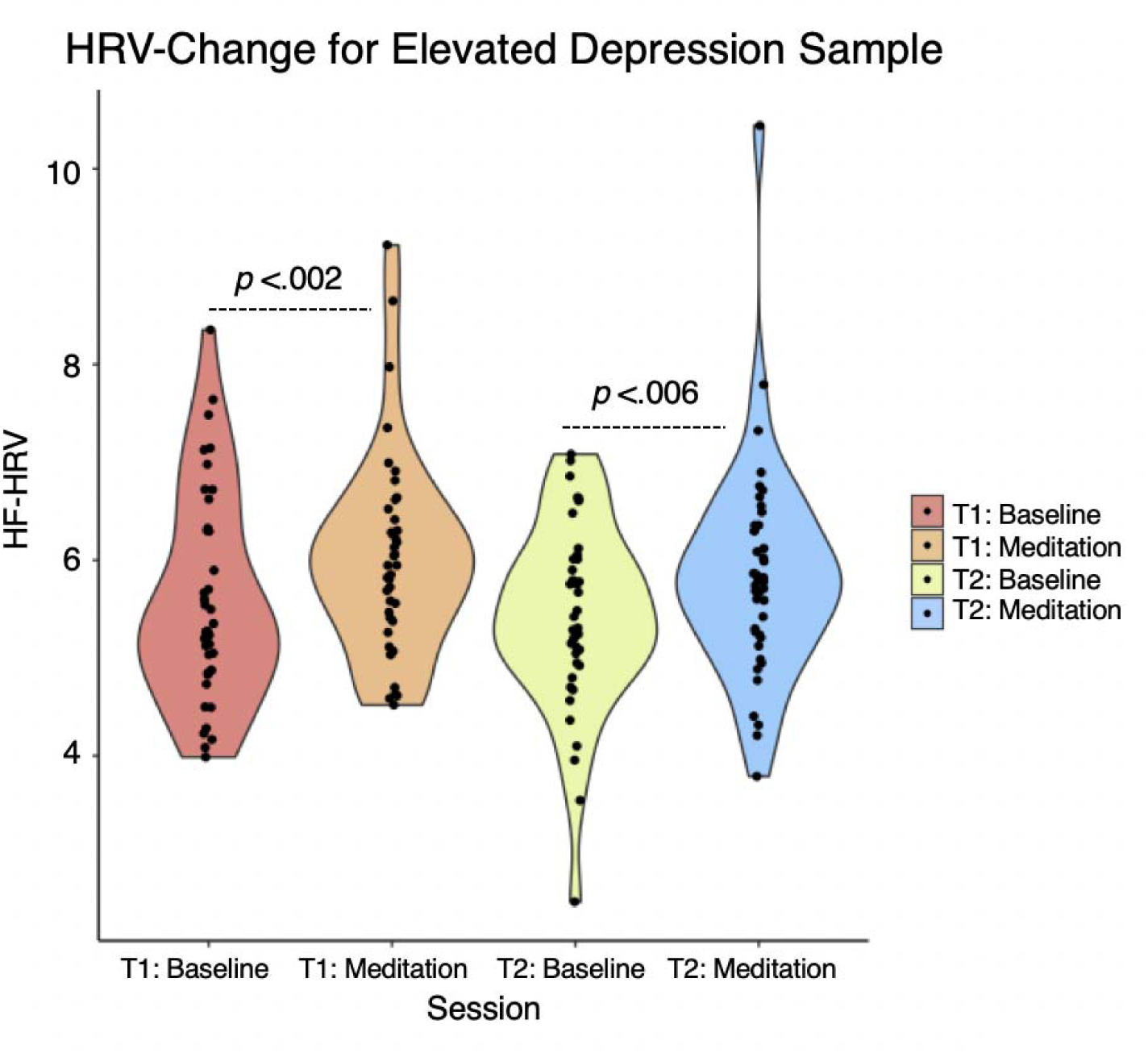
Overall changes in physiology (HRV) derived from our compassion intervention, at rest versus Meditation, at both pre (T1) and post (T2) two-week training. X-axis depicts total 15-minute bins for each session which were transformed to the natural log, and Y-axis indicates subsequent normalized units of HRV.

### RMSSD HRV Response

Convergent evidence for significant HRV shift is also demonstrated when we consider the time-series metric of HRV, RMSSD (i.e., Root Mean Square of Successive Differences). As shown in Figure 3, a two-way ANOVA revealed a significant within-subjects effect of session (i.e., rest vs intervention), whereby HRV was higher when engaged in the compassion intervention than at rest (*F*(1,38) = 15.99, *p* <.001). However, we observed a non-significant within subjects effect of time (i.e., pre and post two-week training) (*F*(1,38) = .31, *p* = .58), whereby rest versus intervention did not differ between timepoints, and a non-significant interaction (*F*(1,38) = 1.15, *p* = .29). Follow-up t-tests revealed significant increases in HRV from T1 rest to T1 intervention (t(39) = 3.115, *p* <.003), as well as from T2 rest to T2 intervention (t(39) = 3.763, *p* <.001). T1 rest to T2 intervention was non-significant (t(40) = 0.667, *p* = .508).

**Figure 3.**
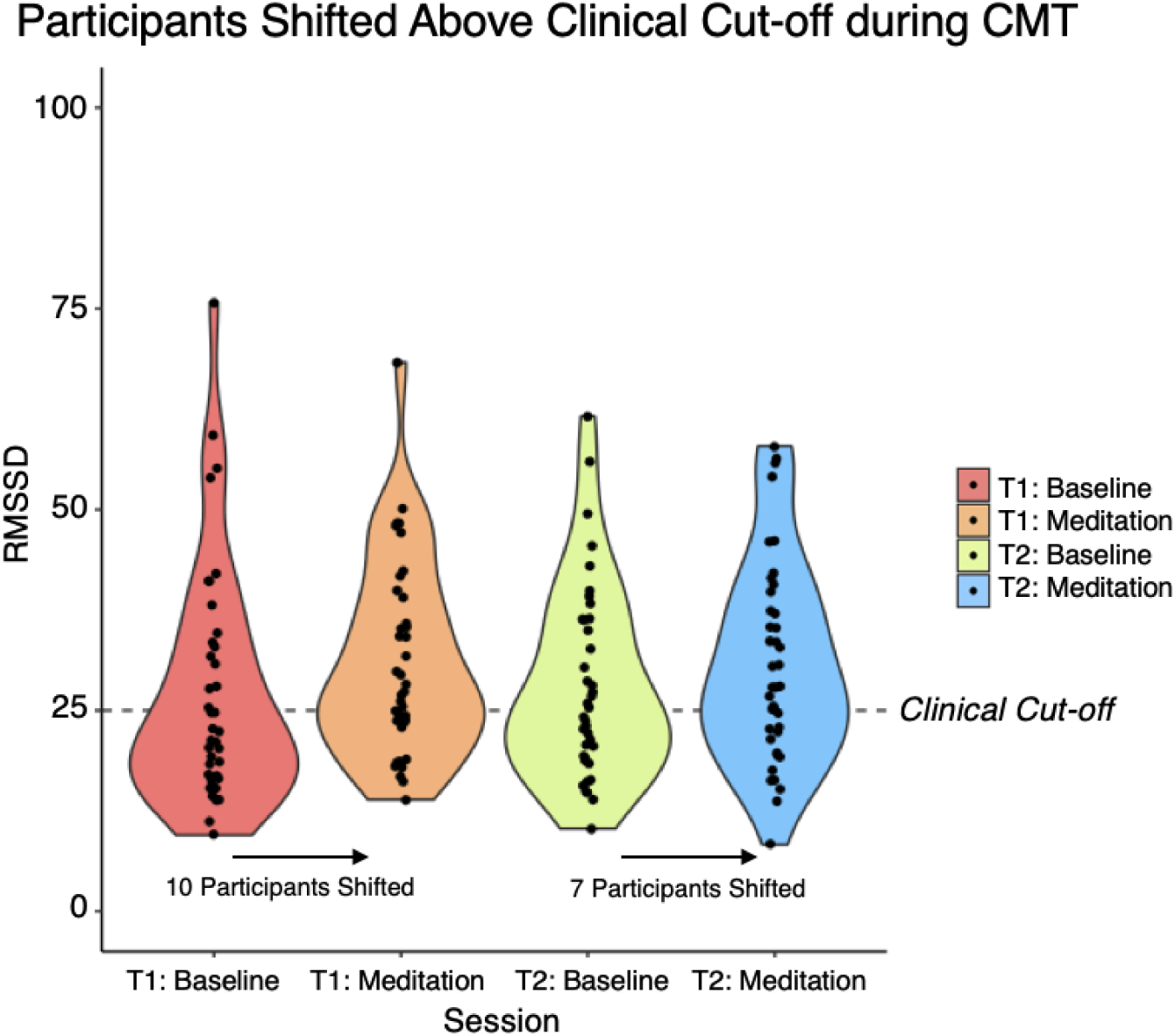
Percentage improvement of a subset of participants shifted out of a clinically at-risk range of poor resting-HRV, after engagement in compassion. X-axis depicts pre- and post-two-week training, at either rest or intervention (compassion meditation), and Y-axis depicts RMSSD HRV value. Dashed-line depicts clinical cut-off (25), N = 41.

### HRV-shift for clinically at-risk participants

Next, we assessed the percentage improvement of participants who were able to shift out of a clinically at-risk range of low resting-HRV, as defined empirically^3^. In accordance with these guidelines, we report raw RMSSD HRV scores for interpretation, and note RMSSD HRV is considered a reliable and valid time-series metric of vagally-mediated HRV^38,40,41^. Here, as also demonstrated in Figure 3, ten and seven clinically-at risk participants were shifted above the clinical cut-off of 25 RMSSD^3^ when engaged in the compassion intervention versus at rest, at either pre- and post-two-week training, respectively.

### Relationship between physiology and behaviour

Next, we examined if Dosage, that is, how often participants engaged in the compassionate practice across the two-week training period, was a significant factor in differential HRV response. To derive a marker of Dosage, we assessed a metric of how often each participant accessed the online recording across the two-week period. Following the approach previously reported in a healthy control sample, we median split this Dosage data to examine if those who “Listened More” versus “Listened Less” was a differential factor in HRV response. However, approximately half of this population did not engage in the exercise across the two weeks (N = 20). Therefore, we chose to examine potential differential HRV response for those who did not engage in the meditation (“No Listens”) versus those who did engage in the meditation (“Listens”). Those who did not listen (N = 20) accessed the recording zero times across the two-week period *(M =* 0, *SD* = 0). In contrast, those who did listen accessed the recording on average 3.9 times across the two-week period (*M* = 4.14, *SD* = 3.87, range = 5-15). A 3-way mixed ANOVA revealed a non-significant between subjects effect, whereby Engagement was not a significant factor in differential HRV response, *F*(1,37) = 0.12, *p* = .0733, *ns* (Figure 4). However, HRV response was overall greater during CFT versus at Rest for both subgroups, *F*(1,37) = 14.69, *p* <.001. We also observed a non-significant effect of session time, whereby HRV response at Time 1 versus Time 2 were not significantly different across the two groups, *F*(1,37) = 1.26, *p* = .269, *ns*).

**Figure 4.**
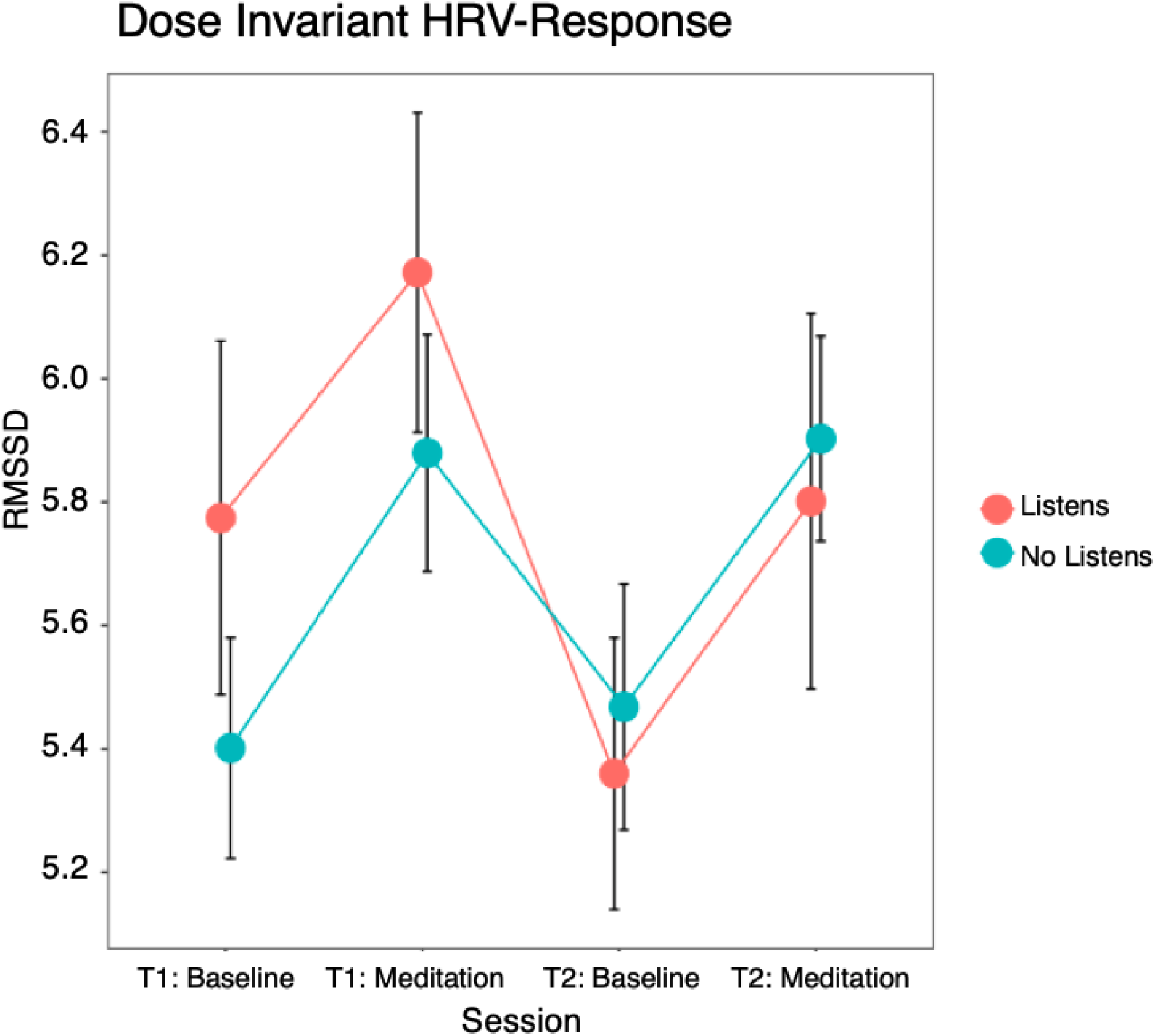
Evidence for dose-invariant Engagement in HRV response. Here we present HRV data across T1 and T2, at Baseline and CFT meditation, plot for participants who Listened (Red Line) versus Did Not Listen (Green Line) to the meditation across the two-weeks. No between-subjects effect across the subgroups were evident.

### 1-minute HRV response

In order to provide greater insight into the moment-to-moment HRV response across these time periods, potentially as a marker of shifting participant’s baseline HRV after two-week training, we visually explored the average HRV signal-change for each participant between Rest and CFT during 1-minute bins, ordered for T1 and T2 (Figure 5). Here, we identified that while HRV baseline at Rest did not increase (or shift) at T2 such as visually shown for a group of heathy control participants within the same paradigm^5^, the HRV recording at T2 during CFT was shown to decrease – potentially as a marker of the threat induction component of the exercise.

**Figure 5.**
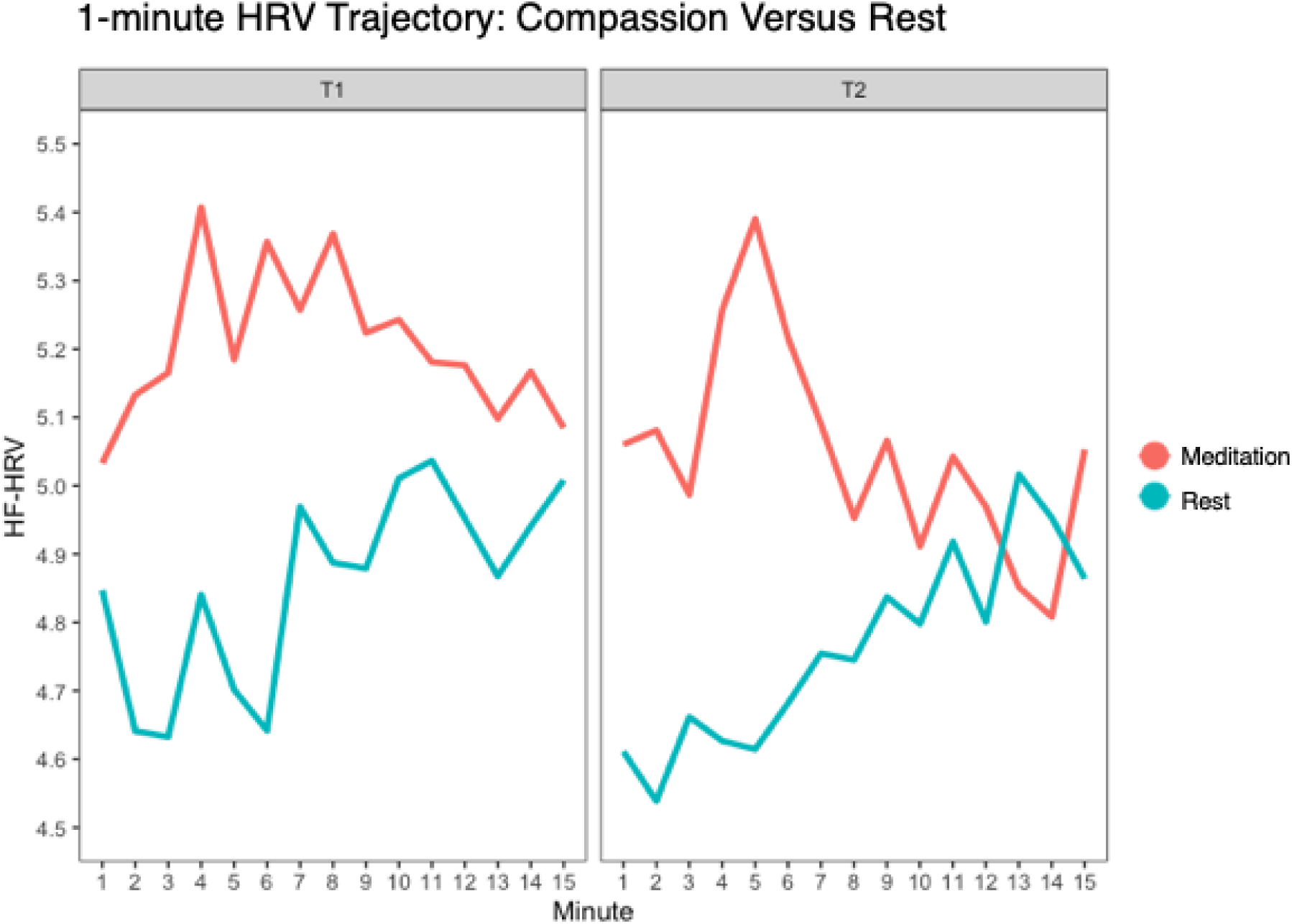
Average HRV signal-change across 1-minute bins, ordered for T1 and T2, at CFT Meditation and Rest. Note the overall between Meditation and Rest at both timepoints, whereas the average HRV recording at 1-minute intervals were greater for the compassion condition (Red Lines) versus the baseline Resting state (Blue Lines). Note that at T2 (right panel), the HRV recording during CFT was shown to decrease during the threat component of the exercise.

## Discussion

The aim of this study was to determine whether a core CFT exercise, ‘the compassionate-self’ could significantly improve HRV for a sample of participants with severe depressive symptoms. Furthermore, we explored whether practice effects (i.e., dosage effect) in CFT across a two-week self-directed training period influenced outcomes. In addition, we assessed whether the CFT practice could shift HRV above a clinical cut-off.

First, we identified significant increases in HRV during CFT versus a resting state at both timepoints (Figure 2,3). Second, we found that HRV response did not change as a function of whether participants practiced the exercise more frequently (i.e., listened more to the compassionate-self exercise) with the self-guided CFT across the two-week training period (Figure 4). Third, we found that CFT was able to ‘shift’ a subset of participants by raising their HRV level above a clinical cut-off of low resting HRV (10 participants at T1 and 7 participants at T2, Figure 3). Fourth, when examining the minute-by-minute HRV signal change during the CFT exercise we demonstrated that while trait-level HRV did not increase after the two-week self-guided training period, HRV during CFT at T2 was shown to decrease across time (Figure 5). A potential explanation for this finding could be a marker of greater engagement in the ‘threat’ component of the CFT exercise – a feature that we discuss below.

Importantly, this work is the first to demonstrate that CFT can significantly increase HRV within a sample of severe depressive symptoms, versus a resting state, indicating how CFT may effectively target the parasympathetic nervous system within both healthy (shown previously)^5,21,23,32,33^ and now depressive populations. This has important clinical implications, as when in session with a depressed client, actively cultivating the compassionate-self will have an immediate positive impact on their HRV activity. This is significant for CFT, as the therapy approach postulates that by engaging in the compassionate-self, the individual will be able to activate a stronger parasympathetic activation, allowing them to be more grounded and work with self-criticism and emotional difficulties from a different motivational system (Kirby & Gilbert, 2017). Our data indicates this is possible at the state level, however, a two-week training period was insufficient at being able to transition this HRV improvement at the trait-level.

While this has implications for therapy and training, it is important to remark on the reduction in HRV during CFT at T2, the poorer engagement in self-guided practice, and the lack of baseline-shift at T2. Indeed, it is possible that the reduction in HRV at T2 is due to greater engagement in the threat of the task, given this component of the exercise invites participants to reflect on a time when they were self-critical of a personal mistake, setback or failure. The self-critical component is introduced at the 11-minute mark, which coincides with a steep drop in the HRV signal (see Figure 5). Thus, this finding echoes previous research that has shown an increase in HRV during compassion engagement, but also a reduction in HRV during threat^21^, a finding observed visually in our data to a greater degree in T2 compared to T1 for the CMT condition (Figure 5).

Indeed, this dynamic HRV response we observed could be reflective of a concept known as HRV reactivity^22,23^. HRV reactivity refers to the dynamic (contextual) nature of the HRV signal, and how it should correspond appropriately to task conditions, such that HRV should increase during periods of soothing and safety^6,22,32^, but also ‘drop’ or decrease during periods of threat and stress^4,21,23,42,43^. Importantly, if HRV does not drop during a period of threat, this may be an insightful manipulation check to infer an individual potentially avoiding the stressful task^23^. To date most research that has examined compassion and HRV have focused on baseline improvements, yet recent exciting work has shown the importance of assessing HRV reactivity toward threat and compassionate action within discrete experimental conditions^21^. Accordingly, it would be insightful for future research to potentially examine specific experimental periods of threat and compassion with a sample elevated in depression, in order to further establish the role of adaptive HRV reactivity with this group.

Importantly, those with severely depressed symptoms can find self-compassion difficult, and can have greater fears of self-compassion^27,44^. Thus, it is not surprising that almost 50% of the sample did not listen to the CFT exercise during the two-week period. The lack of practice with the CFT practice has implications for self-directed delivery of these interventions, as many will avoid the exercise despite the physiological improvements they afford. Novel approaches to help assist with engagement in self-directed delivery of CFT should be considered to help with engagement. For example, with severely depressed groups the implementation of the practice alongside regular contact with a therapist, or telephone support, or even with a reinforcement schedule built into the online delivery (e.g., the participant receives a ‘token’ or ‘badge’ for listening to a track) could improve engagement and dosage. Furthermore, lack of engagement in the exercise could be a candidate explanation for the lack of increase in baseline HRV at T2. It is possible that a longer – and perhaps more intensive – implementation of CFT would be needed to observe a ‘trait’ level increase in baseline HRV within a clinically depressed sample.

Future work can extend on these initial promising findings that CFT can target HRV within groups with severe depressive symptoms by conducting a larger-scale randomized controlled trials. Here, it will be possible to examine how levels of depressive symptomatology or fears of self-compassion may be a significant moderator of HRV during CFT, indicating that these variables will need to be targeted in-order to successfully increase HRV and to derive benefits from CFT. Furthermore, it will be important to explore the timeframe and dosage required of CFT to shift participants above the clinical cut-off of low resting HRV. What is promising, is that CFT can significantly improve HRV for those with severely depressed symptoms at the state-level, and future work should continue to examine CFT’s effectiveness for those with depression.

## Data Availability

All data produced in the present study are available upon reasonable request to the authors

